# Hospitalized COVID-19 Patients treated with Convalescent Plasma in a Mid-size City in the Midwest

**DOI:** 10.1101/2020.06.19.20135830

**Authors:** William Hartman, Aaron S Hess, Joseph P Connor

**Affiliations:** Department of Anesthesiology, University of Wisconsin-Madison; Department of Pathology and Laboratory Medicine, University of Wisconsin-Madison

## Abstract

**Background:** SARS-CoV-2 and its associated disease, COVID-19, has infected over seven million people world-wide, including two million people in the United States. While many people recover from the virus uneventfully, a subset of patients will require hospital admission, some with intensive care needs including intubation, and mechanical ventilation. To date there is no cure and no vaccine is available. Passive immunotherapy by the transfusion of convalescent plasma donated by COVID-19 recovered patients might be an effective option to combat the virus, especially if used early in the course of disease. Here we report our experience of using convalescent plasma at a tertiary care center in a mid-size, midwestern city that did not experience an overwhelming patient surge.

**Methods:** Hospitalized COVID-19 patients categorized as having Severe or Life-Threatening disease according to the Mayo Clinic Emergency Access Protocol were screened, consented, and treated with convalescent plasma collected from local donors recovered from COVID-19 infection. Clinical data and outcomes were collected retrospectively.

**Results:** 31 patients were treated, 16 severe patients and 15 life-threatened patients. Overall mortality was 27% (4/31) but only patients with life-threatening disease died. 94% of transfused patients with severe disease avoided escalation to ICU care and mechanical ventilation. 67% of patients with life-threatening disease were able to be extubated. Most transfused patients had a rapid decrease in their respiratory support requirements on or about day 7 following convalescent plasma transfusion.

**Conclusion:** Our results demonstrate that convalescent plasma is associated with reducing ventilatory requirements in patients with both severe and life-threatening disease, but appears to be most beneficial when administered early in the course of disease when patients meet the criteria for severe illness.

## Introduction

SARS-CoV-2 and its associated disease, COVID-19, has infected over seven million people world-wide, including two million people in the United States. (1) While many people recover from the virus uneventfully, a subset of patients will require hospital admission, some with intensive care needs including intubation, and mechanical ventilation. (2) To date there is no cure and no vaccine is available. In April 2020, the FDA gave permission to the medical community to conduct clinical trials using investigational COVID-19 convalescent plasma (CP) to treat infections in hospitalized patients under a single-patient emergency Investigational New Drug (eIND). (1) Passive immunization with CP was first used as a therapy for diphtheria in 1892 and was the subject for the first Nobel Prize in Physiology and Medicine in 1901. More recently, convalescent plasma was used as a therapy to combat the SARS and MIRS coronaviruses. (4) In March, 2020 the first report of convalescent plasma to treat COVID-19 was published. (3) In the United States the use of convalescent plasma has been made widely available via and FDA approved Expanded Access Protocol (EAP) administrated through Mayo Clinic.

As has been previously described, our institution was one of the earliest donor sites for COVID-19 convalescent plasma in the US, and initially provided 25% of the total national donor pool for the American Red Cross. (5) Our hospital is in Dane County, WI, part of a generous donor community in the Midwest that is a significant net exporter of blood products to the rest of the US. The county population is 546,700 people, half of whom live in the state capital, Madison, where our large, academic tertiary care center is located. Madison is a mid-size city with no high-rise buildings, a modest public transit / bus system and a median income of $71,790.00. It is also home to the state government and the flagship University of Wisconsin campus, which has 44,000 students including many national and international enrollees. While larger cities like New York City and Seattle were experiencing large surges in the volume of critically ill patients in their health systems, the health care system in Madison saw relatively modest numbers of COVID-19 patients. (8-10)

We describe our experience treating a series of COVID-19 patients with severe or life-threatening disease with CP. Ventilatory and final patient outcomes are discussed, and the results compared to previously published data on CP treatment.

## Methods

This study was approved by the University of Wisconsin Institutional Review Board. Cases met all criteria for enrollment under the Mayo Clinic Expanded Access Protocol (IND # 20-003312) and gave written, informed consent for CP transfusion and data collection. Enrollment criteria are described elsewhere. (11) Briefly, all enrollees had laboratory confirmed COVID-19 with either severe or life-threatening disease. Severe disease was defined as the presence of subjective dyspnea, a respiratory frequency ≥ 30/min, blood oxygen saturation ≤ 93% on room air, partial pressure of arterial oxygen to fraction of inspired oxygen ratio < 300, and/or lung infiltrates >50% within 24 to 48 hours. Life-threatening disease was defined as respiratory failure, septic shock, and/or multiple organ dysfunction or failure at the time of transfusion.

COVID-19 convalescent plasma was collected from a local donor recruitment and referral program in collaboration with the American Red Cross. Briefly, in response to guidance from the FDA dated April 3, 2020 We convened a local working group to establish a University of Wisconsin Hospital-based COVID-19 Convalescent Plasma program for both candidate recipients and potential COVID-19 recovered donors. Stakeholders were assigned to issues within their expertise including transfusion, the University’s Office of Clinical Trials (OCT), the Media Relations, and the local American Red Cross (ARC) donor center. The Transfusion Medicine section developed an inventory and ordering process for CCP units within our electronic medical record system. The OCT worked closely with ARC and FDA to ensure compliance with rapidly evolving rules. OCT staff also worked with potential donors identified by UW clinician referral or self-referral through local media coverage and via information provided to patients in the discharge instructions after all COVID-19 related hospital admissions. Potential donors were then screened and recruited to donate via a scripted telephone interview. The local ARC established a process for receiving prospective donors and worked with national leadership to develop long-term protocols.

Recipient data were abstracted from the medical record into a standardized case report form. Results were presented with descriptive statistics. Parametric and non-parametric tests were used as appropriate. All analyses were performed using commercially available statistical software (Enterprise Guide 7.1, SAS, Cary, NC).

## Results

All appropriate regulatory, IRB and patient care protocols were in place to begin transfusions by April 9, 2020. The first unit from our donor screening program was collected on April 10, 2020 with acceptable units available for patient transfusion on April 12, 2020.

Between April 12 and June 2, 2020, there were 62 inpatient admissions for COVID-19 with severe or life-threatening disease. Of these, 31 met all inclusion criteria, gave consent, and received convalescent plasma. Among the 31 transfused patients, 15/31 (48%) met criteria for life-threatening disease, and 16/31 (52%) met criteria for severe disease at time of transfusion. Five patients were already in-house and were consented to receive plasma before units were available from our blood supplier. Four of these five patients had already progressed to or had presented with life-threatening disease at the time of transfusion. Characteristics of transfused patients are summarized in Table 1. In general, treated patients were not severely ill as measured by Sequential Organ Failure Assessment (SOFA) scores, but inflammatory markers were elevated. A comparison of characteristics between transfused patients graded with severe or life-threatening illness are summarized in Table 2. Patients with life-threatening disease were not significantly different from those with severe disease at time of transfusion with regard to sex, BMI, d-dimer or ferritin levels, but had significantly higher SOFA scores and c-reactive protein levels and had longer hospital stays. No patients with severe disease died, compared to four (27%) with life-threatening disease (p = 0.032).

**Table 1:** Characteristics of patients receiving COVID-19 convalescent plasma at time of transfusion. *

| Characteristic | All patients<br>(N = 31) |
| --- | --- |
| Sex – no. (%) |  |
| Female | 10 (32) |
| Male | 21 (68) |
| BMI | 37.4 ± 10.5 |
| Classification of COVID-19 Illness – no. (%) |  |
| Severe | 16 (52) |
| Life-threatening | 15 (48) |
| Respiratory support at time of transfusion – no. (%) |  |
| None | 2 (6) |
| Low-flow nasal cannula | 11 (35) |
| High-flow nasal cannula | 8 (26) |
| Mechanical ventilation | 10 (32) |
| Sequential Organ Failure Assessment (SOFA) Score † | 2 [4] |
| C-reactive protein – mg/dL | 15.5 ± 9.5 |
| D-dimer – mcg/mL | 2.6 ± 4.0 |
| Ferritin – ng/mL | 1532 ± 1414 |
| Hospital length of stay in days | 12 [22] |
| Days from CP transfusion to discharge | 7 [14] |
| Final disposition – no. (%) |  |
| Home | 15 (48%) |
| Inpatient rehabilitation then home | 2 (7%) |
| Skilled nursing facility or long-term care | 5 (5%) |
| Dead | 4 (13%) |
| Ongoing inpatient care | 5 (16%) |

\*Plus-minus values are means ± SD, values with brackets are medians [interquartile range].

**Table 2:**
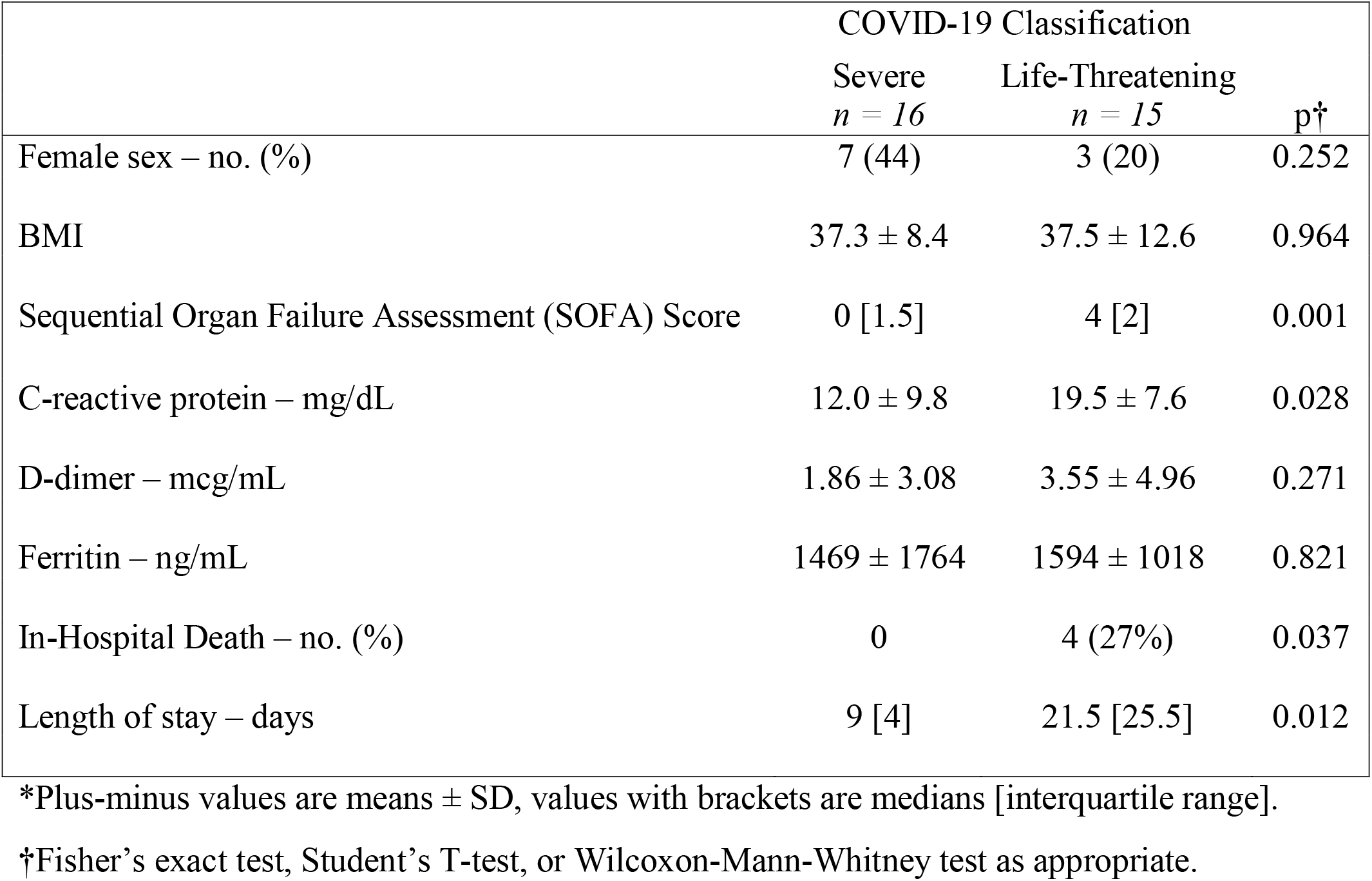
Characteristics of patients with severe vs. life-threatening disease receiving COVID-19 convalescent plasma at time of transfusion. *

Inpatient respiratory support requirements over time for patients with severe disease are summarized in Figure 1. Among the 16 patients that were transfused for severe disease one (6%) had progressive respiratory dysfunction and ultimately required intubation five days after transfusion of convalescent plasma (eight days after hospital admission, 13 days after onset of symptoms). Another remains inpatient on room air with persistently positive SARS-CoV-2 PCR testing and is awaiting transfer to a skilled nursing facility. Of the remaining patients with severe illness, all fourteen were discharged, most on room air. Three of these patients were transferred to skilled nursing or long-term care facilities and the remainder went home with self-care. The median length of hospitalization in this groups was 9 days (mean 11.1 ± 6.9 days and range 4-29 days).

**Figure 1:**
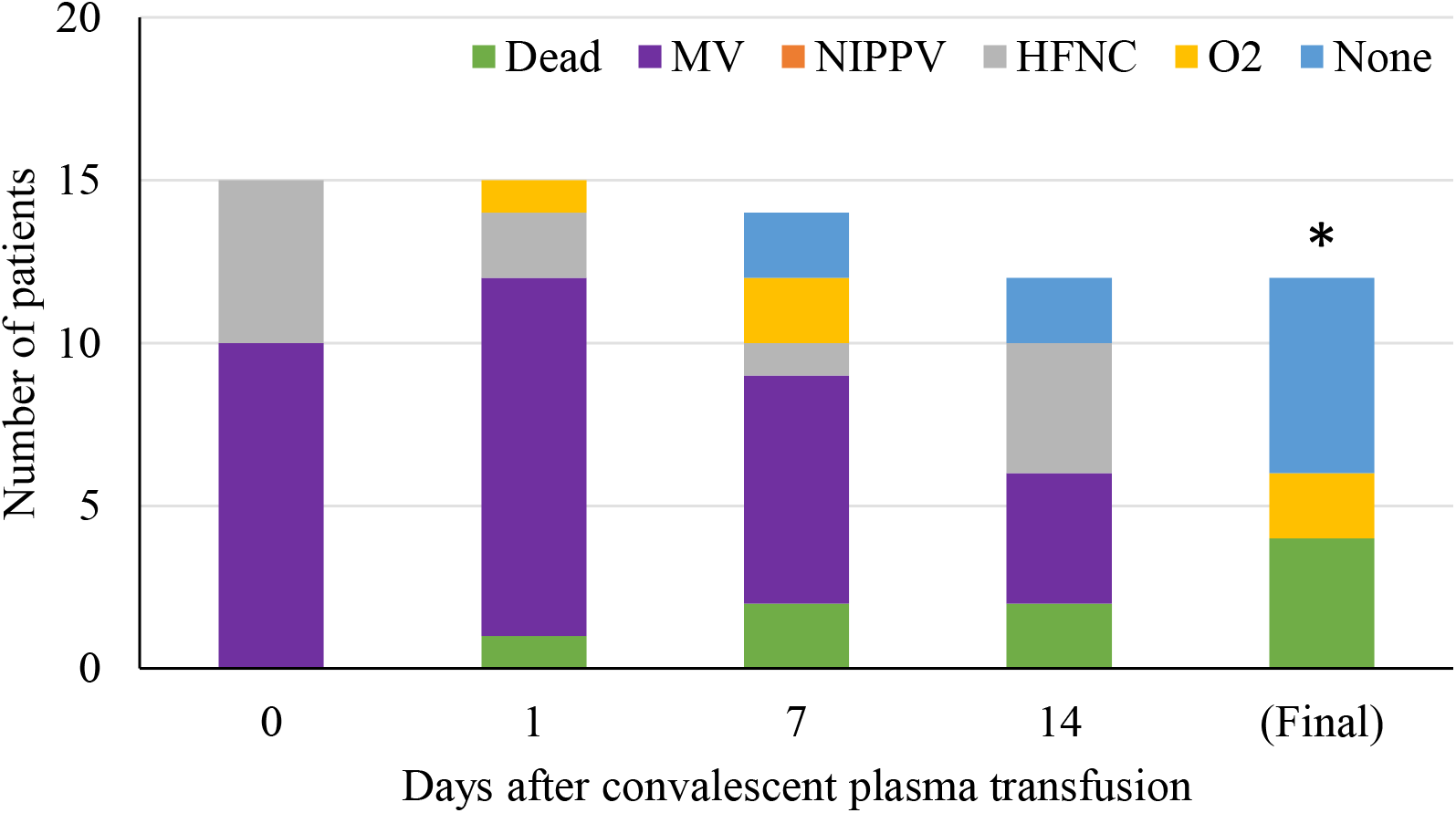
Inpatient respiratory support type by hospital day among COVID-19 patients with life-threatening disease receiving convalescent plasma (n = 15). The asterisk (*) marks that three patients were excluded from the tally because their final respiratory status is not known. At the time of last follow-up, two were on mechanical ventilation and one was on high-flow nasal cannula.

Inpatient respiratory support requirements over time for patients with life-threatening disease are summarized in Figure 2. Twelve (80%) required intubation and mechanical ventilation at some point during their hospital stay. Nine of these were intubated for management of acute respiratory failure within 24 hours of admission to the hospital, the other 3 were intubated for progressive respiratory failure 5, 5, and 7 days after admission respectively. Only one of the 12 ventilated patients received CP prior to intubation: he was on 100% O2 via high-flow nasal cannula at the time of transfusion and was intubated 15 hours later. Ultimately, 8 (67%) were extubated, after a median of 10 days since CP transfusion. Six of the eight extubated patients have been discharged from the hospital: four to self-care at home, one to a skilled nursing facility, and one to a long-term care facility. The median length of stay for the six patients that required mechanical ventilation (excluding the patients that died or remained hospitalized) was 26 days (IQR 19.75, range 5-38 days).

**Figure 2:**
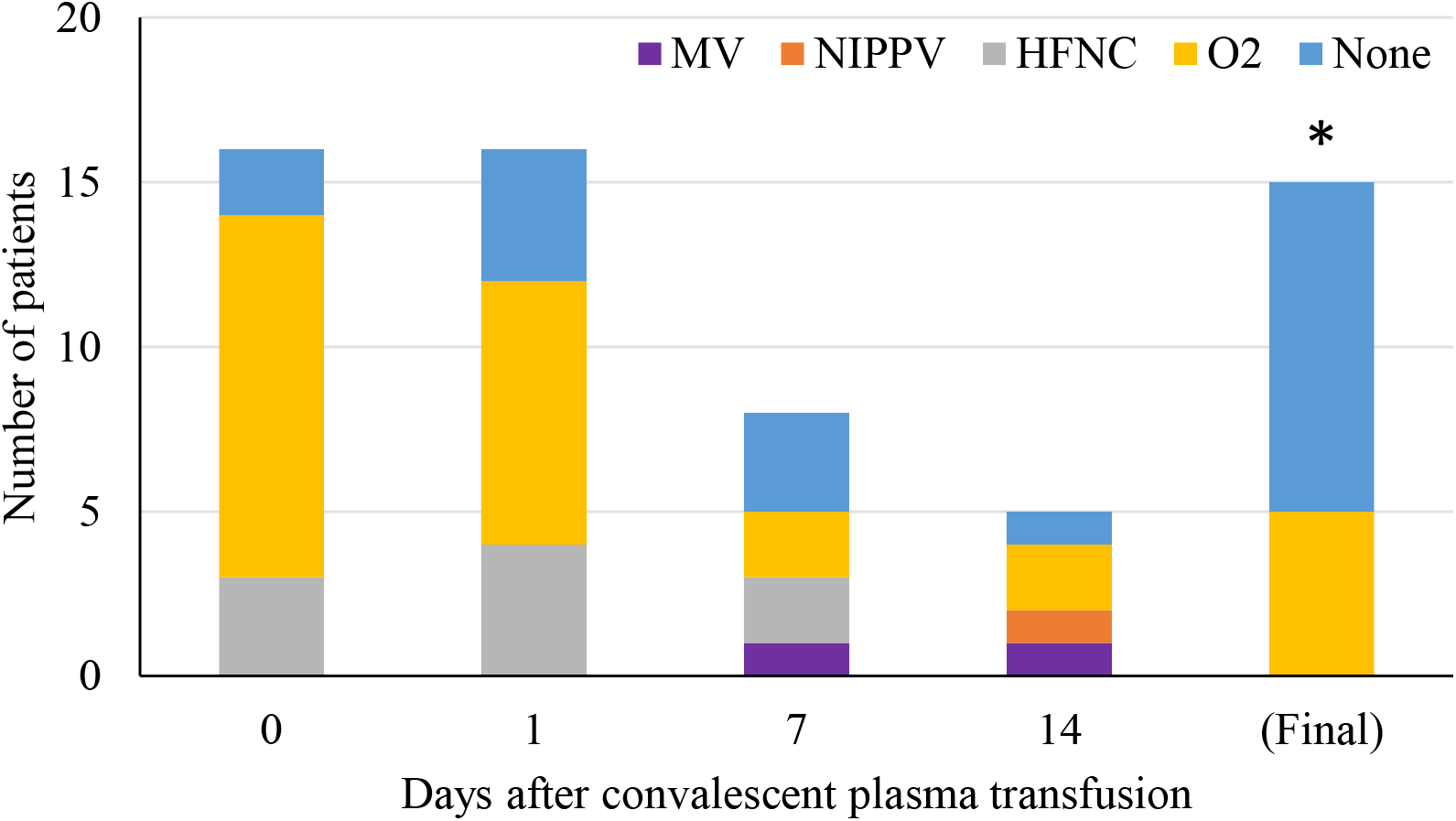
Inpatient respiratory support type by hospital day among COVID-19 patients with severe disease receiving convalescent plasma (n = 16). The asterisk (*) marks that two patients were excluded from the tally because their final respiratory status was not known. At the time of last follow-up, one was on mechanical ventilation and one was on low-flow nasal cannula.

Of the three patients with life-threatening disease who were not intubated at the time of transfusion, all three were in severe respiratory distress. One was transferred from a memory care facility and had a signed Do Not Intubate order, one had recently self-extubated during an episode of delirium and did not require re-intubation, and one adamantly refused intubation.

## Discussion

We report a case series of 31 patients with either serious or life-threatening COVID-19 infection treated with COVID-19 CP who demonstrated favorable clinical outcomes when compared to those reported in the literature to date. For both severe and life-threatened patients, respiratory support requirements began to decrease at about day 7. Once the ventilatory requirements began to decrease, they did so rapidly. Most patients were able to be discharged home on room air. The overall mortality was 13% (4/31). Among patients who were admitted with infection that met the criteria for severe disease and were transfused CP prior to the development of respiratory failure the mortality to date is zero, and only one patient (6%) has had subsequent escalation of respiratory failure requiring mechanical ventilation. Among our patients with life-threatening disease we report an extubation rate of 67%. Overall, compared to outcomes reported in the literature to date, patients transfused CP appear to have better outcomes in the face of both severe and life-threatening disease. (12,13)

Our results are consistent with other early reports of outcomes in COVID-19 patients transfused with CP. A recent cohort study by Liu and colleagues of 39 cases and 156 matched controls from Mount Sinai hospital in New York City reported a 12.8% mortality rate among patients with severe or worse disease who received convalescent plasma, and significantly better outcomes among patients transfused prior to mechanical ventilation. (14) Transfused patients were very similar to ours with respect to inflammatory markers and distribution of respiratory support requirements. Although we have no basis for internal comparison because our transfused rate was high and overall case numbers prevented us from matching a control group, our experience reinforces the suggestion that early administration is of greater clinical benefit than delaying transfusion under the development of severe disease. This is in line with one of the first published randomized clinical trials of CP, in which Li and colleagues found clinical improvement was limited to those with without life-threatening disease, with 91% improvement in the plasma group compared to 68% in the control arm. (15)

We observed a high rate clinical improvement among mechanically ventilated patients who received COVID-19 CP. Although 4 (29%) of our patients with life-threatening disease died, 9 (64%) has improve respiratory by 14 days after transfusion. Improvement with convalescent plasma in patients already requiring mechanical ventilation is in line with one of the early reports of convalescent plasma treatment by Shen et al., who reported on improvement in multiple clinical parameters in five of five (100%) patients transfused convalescent plasma. Li and colleagues, by contrast, saw only a 10% rate of improvement among intubated patients receiving CP 14 days after transfusion.

One potential confounding factor in the improved outcomes we have seen could be the regional/geographical differences in outcomes as have been reported in the literature. Unlike the large patient surges experienced in Seattle and then in New York City, the healthcare system in Dane County/Madison, including at University Hospital, has not experienced a large or overwhelming surge of patients. In the initial reports from the Seattle area, a substantial number of patients (81%) were initially admitted to the intensive care unit, requiring intubation and mechanical ventilation, and by mid-March, 2020, they reported a mortality of 67% and the continued critical care needs of an additional 24% of patients. (10) Similarly, from March and April, 2020, New York City area hospitals experienced mortality rates as high as 70 to 90% in patients requiring mechanical ventilation. (11) Among 257 critical care admissions in the New York Presbyterian/Columbia hospital system 39% have died with the death rate being much higher at 79% for the 203 patients requiring mechanical ventilation. By contrast the overall death rate in our transfused cases was only 13% and to date, at worst 29% in those requiring mechanical ventilation. (12)

Ji and colleagues described how infection prevalence compared to resource availability and healthcare burden are directly related to differences in mortality as seen in various areas of China. For instance, in the Wuhan province, the epicenter of the pandemic, mortality rates were reported to be greater than 3-4% where in other, even more populous areas, of China mortality was less than 1%. They go on to describe that mortality appears to be related to the incidence of disease per capita and the resources available in the local healthcare system to absorb and care for the number of cases in need. (16) This may well be part of the phenomenon of less aggressive appearing disease seen in the Dane County and Madison area. Dane County, with a population of approximately 550,000, has an infection rate of 143/100,000 population and a mortality rate of only 5/100,000. Other more populated and dense areas such as New York City have incidence rates 20 times greater than ours and mortality rates 50 times higher. (12)

Our report has significant limitations. Because our institution did not see a large COVID-19 patient surge like those experienced elsewhere, we were unable to develop a well-matched control group for the purposes of comparison. Our patient population is also relatively homogenous and with good access to medical care. In addition, because most of our CP came from donors in the earliest phases of collection, we did not have donor antibody titers available for analysis.

In summary, our experience to date at a large Midwest academic medical center demonstrates evidence of clinical benefit for patients treated with COVID-19 convalescent plasma in both the severe and life-threatening patient populations. Although our data supports the promise of this therapy it also underscores the importance of ongoing and additional planned randomized clinical trials of CCP in the full spectrum of COVID-19 clinical presentations from salvage therapy in critically ill, mechanically ventilated patients to post-exposure prophylaxis/prevention in high risk individuals. (17)

## Data Availability

Data is not currently available.

## Acknowledgements

We are grateful for the tremendous help in preparing this manuscript by the University of Wisconsin-Madison Office of Clinical Trials.

Funding provided by the University of Wisconsin-Madison Institute of Clinical and Translational Research (ICTR).

This study was supported in part by a US Department of Health and Human Services (HHS), Biomedical Advanced Research and Development Authority (BARDA) grant contract 75A50120C00096, National Center for Advancing Translational Sciences (NCATS) grant UL1TR002377, Schwab Charitable Fund (Eric E Schmidt, Wendy Schmidt donors), United Health Group, National Basketball Association (NBA), Millennium Pharmaceuticals, Octopharma Octapharma USA, Inc, and the Mayo Clinic.

